# Stroke Severity, Caregiver Feedback, and Cognition in the REGARDS-CARES Study

**DOI:** 10.1101/2023.10.26.23297649

**Authors:** Jason A. Blake, D. Leann Long, Amy J. Knight, Burel R. Goodin, Michael Crowe, Suzanne E. Judd, J. David Rhodes, David L. Roth, Olivio J. Clay

## Abstract

**Objective:** Cognitive impairment after stroke is common, present up to 60% of survivors. Stroke severity, indicated by both volume and location, is the most consequential predictor of cognitive impairment, with severe strokes predicting higher chances of cognitive impairment. The current investigation examines the associations of two stroke severity ratings and a caregiver-report of post-stroke functioning with longitudinal cognitive outcomes.

**Methods:** The analysis was conducted on 157 caregivers and stroke survivor dyads who participated in the Caring for Adults Recovering from the Effects of Stroke (CARES) project, an ancillary study of the REasons for Geographic and Racial Differences in Stroke (REGARDS) national cohort study. Glasgow Outcome Scale (GOS) and modified Rankin Scale (mRS) collected at hospitalization discharge were included as two primary predictors of cognitive impairment. The number of caregiver-reported problems and impairments at nine months following stroke were included as a third predictor. Cognition was assessed using a biennial telephone battery, incorporating multiple cognitive assessments to assess learning, memory, and executive functioning. Longitudinal cognitive scores were analyzed up to five years post-stroke, controlling for baseline (pre-stroke) cognitive scores and demographic variables of each stroke survivor collected at CARES baseline.

**Results:** Separate mixed models showed significant main effects of GOS (b=0.3280, p=0.0009), mRS (b=-0.2119, p=0.0002), and caregiver-reported impairments (b=-0.0671, p<0.0001) on longitudinal cognitive scores. In a combined model including all three predictors, only caregiver-reported problems significantly predicted cognitive outcomes (b=-0.0480, p<0.0001).

**Impact:** These findings underscore the importance of incorporating caregivers feedback in understanding cognitive consequences of stroke.

## INTRODUCTION

Stroke is a leading cause of death and disability globally.^1, 2^ The prevalence of post-stroke cognitive impairment (PSCI) is present in up to 60% of stroke survivors following their first stroke.^3, 4^ PSCI can be difficult to measure as stroke recovery is a highly variable and individualized process that does not adhere to a fixed timeline.^5^ The first six months after a stroke, termed the “critical recovery window,” are characterized by the most dramatic improvements in the physical and cognitive functioning of the survivior.^6^ However, the degree of these improvements greatly differs among stroke survivors after six months; some experience vast recovery, leading to fewer residual deficits, while others may show more modest progress. The survivors who experience larger functional improvements during this critical period are more likely to demonstrate better cognitive scores in subsequent years.^7, 8^ This variable rate of recovery can contribute to a broad range of cognitive and functional outcomes in post-stroke recovery.

To better improve the outcomes of stroke survivors, it is necessary to understand factors that may be prognostic of post-stroke cognitive recovery. Presence of functional impairment due to stroke severity has been shown to predict worse long-term functional outcomes^9, 10^ including occupational^11, 12^, depressive or other psychosocial symptoms^13, 14^, and overall quality of life^15, 16^ in survivors. In previous studies, it has been demonstrated that initial stroke severity, indicated by both volume and location of stroke, is the most consequential predictor of cognitive impairment, with severe strokes predicting higher chances of cognitive impairment.^17, 18^

Stroke severity is often determined through an objective evaluation of neurological functioning at the time of hospitalization and at discharge. An objective evaluation of acute post-stroke functioning is the Glasgow Outcome Scale (GOS) which is used to assess the broad degree of disability following hospitalization of an acquired brain injury, such as stroke.^19^ Severity is also commonly determined through an assessment of the patient’s functional status that measures capacity to carry out activities of daily living (ADLs), one such functional assessment of stroke is the modified Rankin Scale (mRS).^20^ These types of outcome measures play important roles assisting with clinical decision-making, treatment planning, and outcome assessment. However, these injury measures, including the commonly used National Institute of Health Stroke Scale (NIHSS), have limitations in their ability to predict functional outcomes.^21, 22^ These limitations become especially evident with subtle cognitive or physical changes after stroke, especially when the patient is unaware of their own functioning. In these cases, it is often necessary for clinicians to involve an informant who can provide additional information about the patient’s functional status.^22, 23^

Caregivers, who spend the most time with stroke survivors, are in a unique position to provide valuable insight into the survivors’ functioning. Their observations often complement clinically administered assessments and contribute to a more comprehensive understanding of stroke survivors’ cognitive challenges and recovery process.^24^ In the Alzheimer’s disease and dementia population, the concerns of the patient’s informal caregivers become invaluable as patients often deny cognitive changes in the early stages.^25^ Clinicians also rely on caregiver report when identifying the efficacy of a new medications or treatment used for dementia patients.^26^ Collecting caregiver or informant reports during a standard neuropsychological assessment is one of the vital components for identifying changes in baseline cognition while diagnosing vascular cognitive impairment and dementia.^27^

For this investigation, it is hypothesized that more severe stroke, as measured by clinically administered scales at hospital discharge, is predictive of lower longitudinal cognitive scores in stroke survivors. Additionally, a higher number of impairments and problems reported by the caregivers is hypothesized to be associated with lower cognitive scores in stroke survivors over time. The inclusion of caregiver reports, particularly after the six-month critical recovery window, may provide a more comprehensive picture of the patient’s cognitive and functional status and improve the accuracy of predictions of long-term outcomes following a stroke.

## METHODS

### Participant Recruitment

All participants were recruited via the REasons for Geographic And Racial Differences in Stroke (REGARDS) study. REGARDS is an ongoing national epidemiologic cohort study investigating stroke incidence and mortality that initially enrolled 30,239 participants from 2003 to 2007^28^. The REGARDS study enrolled participants aged 45 or older and included 30% from the Stroke Belt (encompassing Southeastern U.S. states), 20% from the Stroke Buckle (a subset of the Stroke Belt, specifically the coastal plain region of North Carolina, South Carolina, and Georgia), and the remainder from elsewhere in the continental U.S. Each region maintained a balanced demographic, with roughly equal representation of White and Black individuals, and a nearly equal gender distribution within each race-region group. The REGARDS study intentionally oversampled Black individuals due to the study’s specific interest in the comparison of those two racial groups. Once enrolled in REGARDS, participants were administered bi-annual cognitive testing over the phone to measure several cognitive domains such as memory and verbal fluency/executive functioning. All relevant measures and assessments are detailed below. The methodologies for participant sampling, recruitment, and data collection in the REGARDS study have been described further in prior studies.^28–30^ Due to the sensitive nature of the data collected for this study, requests to access the dataset from qualified researchers trained in human subject confidentiality protocols may be sent to REGARDS at.

### CARES Enrollment

Stroke survivors that were initially enrolled in the REGARDS project were then recruited, along with their primary caregivers, to participate in the ancillary project CARES (Caring for Adults Recovering from the Effects of Stroke). Study enrollment for CARES occurred from 2005 to 2016. Eligibility for the CARES study required stroke survivors to (1) be living in the community nine months post-stroke, excluding those in nursing homes or other institutional settings; (2) have a family member or close friend acting as an informal caregiver, or had one immediately following stroke; and (3) have the caregiver also agree to participate in the CARES project.^31^ Once participants were enrolled in the CARES study, the stroke survivor and caregiver dyads were administered the initial baseline CARES interview.^32^ The CARES interviews were conducted via telephone approximately nine months following the stroke event. Of the stroke survivor group, all self-reported a first-time stroke during a follow-up call. The survivors’ medical records were also examined independently to collect measures of stroke severity at the time of hospitalization discharge, confirm a stroke event, and determine the type of stroke when possible. Roth, et al. ^31^ provides a detailed overview of the data collection procedures employed in the CARES study. Both projects were reviewed and approved by the Institutional Review Boards of the University of Alabama at Birmingham and each participating institution.

### Demographics

Demographic information was collected from both the stroke survivors and their caregivers during their enrollment into the REGARDS study as well as during the initial CARES interview. The stroke survivor and caregiver demographic variables used in this analysis, including race, educational level, age, and gender, were collected at CARES enrollment. Sex (male or female), race (Black/AA or White) and marital status (married or unmarried) were dichotomous variables based on self-report.

### Measures

#### Stroke Severity

GOS^33^ and mRS^34^ scores were collected from hospital chart abstraction at time of discharge. The GOS and mRS are commonly utilized in clinical settings as measures of acute stroke recovery and stroke severity. The GOS categorizes injury outcomes into five broad categories: death, persistent vegetative state, severe disability, moderate disability, and good recovery. Possible scores on the GOS range from one, indicating death, to five, indicating good recovery.^19^ As a measure of functional ability, the mRS assesses the degree of disability or dependence in daily activities. The mRS offers a broader perspective on stroke-related functional limitations, with possible scores ranging from zero, indicating no symptoms, to six, representing severe disability or death.^34^

Each caregiver reported the total number of stroke-related impairments at CARES enrollment, an average of nine months after stroke. This 28-item instrument collected caregiver-reported problems (CGRP) that interfered with their activities of daily life (ADL) and instrumental activities of daily living (IADL) over the previous week. The instrument items were drawn from commonly used instruments including the Frenchay Activities Index ^35^, Barthel index^36^, and the Revised Memory and Behavior Problems Checklist.^37^ This measure was used in a previous CARES analysis to determine severity of stroke impairments and problems^38^ and has shown good concurrent validity (correlating over .62 with the Barthel Index and Rankin scores) with observer-assessed indicators of cognitive impairment in stroke.^39^ In previous analysis, coefficient alpha for the scale was 0.92.^39^

#### Cognitive Assessment

The cognitive assessment is a brief telephone battery that is administered to each REGARDS participant every two years. The intervals for collection of this battery are less frequent to lower participant burden. This battery includes the following six tasks to assess the cognitive domains of learning, memory, and executive functioning: a) Word List Learning (WLL) and Delayed Recall (WLDR), from the Consortium to Establish a Registry for Alzheimer’s Disease (CERAD) battery^40^; b) Animal (Semantic) Fluency; c) Letter-F (Phonemic) Fluency; and d) Registration, recall, and orientation from the Montreal Cognitive Assessment (MoCA)^41^ as a part of the National Institute of Neurological Disorders and Stroke - Canadian Stroke Network Vascular Cognitive Impairment Harmonization Standards (NINDS-CSN) five-minute Battery.^42^

The WLL task includes three learning trials of ten-word list, followed by WLDR after a 5-minute delay. Correct responses on the WLL were summed across all three trials and final scores ranged from zero to thirty. On the WLDR, participants recalled as many of the ten-word list as possible, with scores ranging from zero to ten. From the NINDS-CSN, scores for the five-item registration range from zero to five, scores on the five-item recall range from zero to five, and scores on the six-item orientation range from zero to six. Letter fluency prompted participants to name as many words that begin with the letter “F” as they can in one minute. Semantic fluency prompted participants to name as many animals as they could in one minute. Scores on both fluency measures included the total number of valid responses produced by each participant within sixty seconds, after subtracting repetition and intrusion errors. The measures used in this battery are further outlined in the appendix of a previous REGARDS manuscript by Wadley, et al. ^43^ These measures have been validated for reliable administration using telephone interviews.^44^ The raw scores for each assessment have been normed to z-scores according to the patient’s age and sex. The cognitive composite score, the average of the z-scores at each assessment, served as the outcome variable.

#### Statistical Analysis

The data analyses for this paper were performed using SAS^©^ software Studio version 3.81.^45^ Descriptive statistics were reported for the participant dyads included means and standard deviations for continuous measures and percentages for categorical variables. Age, gender, race, and education for each stroke survivor were collected at the CARES baseline interview and included as covariates in all models. P values < 0.05 were considered statistically significant.

The three predictor variables utilized in this analysis are GOS scores, mRS scores, and total number of stroke-related problems as reported by their caregiver. This analysis included the most recent pre-stroke evaluation for each participant as a measure of baseline cognitive functioning. To assess the relationship between post-stroke functional ability and long-term cognitive trajectories, separate mixed effects regression models were used for each predictor. A combined model including all three predictors was also included in this analysis to assess the independent predictive ability of each measure in the presence of the other predictors. These models were structured to examine logarithmic trajectories of cognition over time (number of years post-stroke). Logarithmic trajectories are recommended over linear trajectories when assessing longitudinal cognitive scores in stroke survivors^46, 47^. Each mixed model included in this analysis were covariate adjusted for pre-stroke cognitive scores, time after stroke (years), age, race, sex, and education. Timepoints greater than five years were removed to minimize survivorship bias. Random intercepts and slopes for time at the subject level were included with an unstructured covariance matrix.

Figures 1-3 display the logarithmic relationship between the three measures of stroke severity and cognitive composite scores over time. Participants were divided into two groups based on the 50th percentile (median) of each predictor as a cut-off point. The division of participants into two groups based on the median score allows for a visualization of contrasting cognitive outcomes when grouped by severity of stroke symptoms as classified by each predictor. Least squares means were used to calculate the predicted cognitive outcomes. In all three figures, the blue group represents lower stroke severity, as determined by the assigned predictor variable, while the red group represents worse stroke severity. The model for each figure includes baseline cognition, age at CARES enrollment, gender, race, and education as covariates. As these models use the natural logarithm of time, the first timepoint is estimated at 0.01 years (∼4 days) post stroke. All p-values and statistical significances are based on 95% confidence intervals.

**Figure 1.**
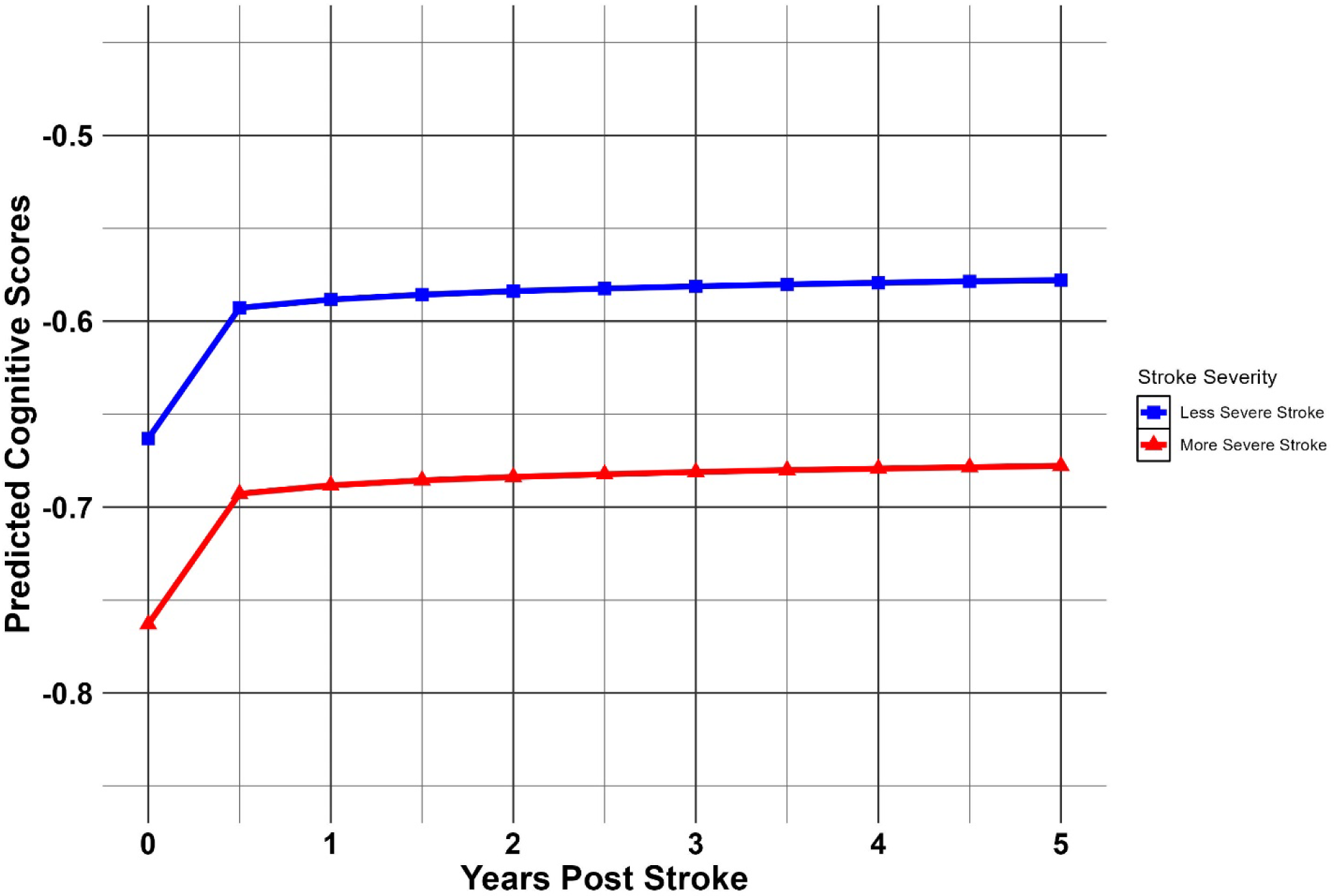
Covariate Adjusted, Logarithmic Trajectories of Cognitive Scores by GOS. The cutoff for ‘more severe stroke’ included any participants with GOS scores less than the median value (5.0). Cutoff for ‘less severe stroke’ were scores greater than or equal to the median value. This model includes baseline cognition, age at enrollment, gender, race, and education as covariates. As this model uses logarithm of time, the first timepoint is estimated at 0.01 years (∼4 days) post stroke.

**Figure 2.**
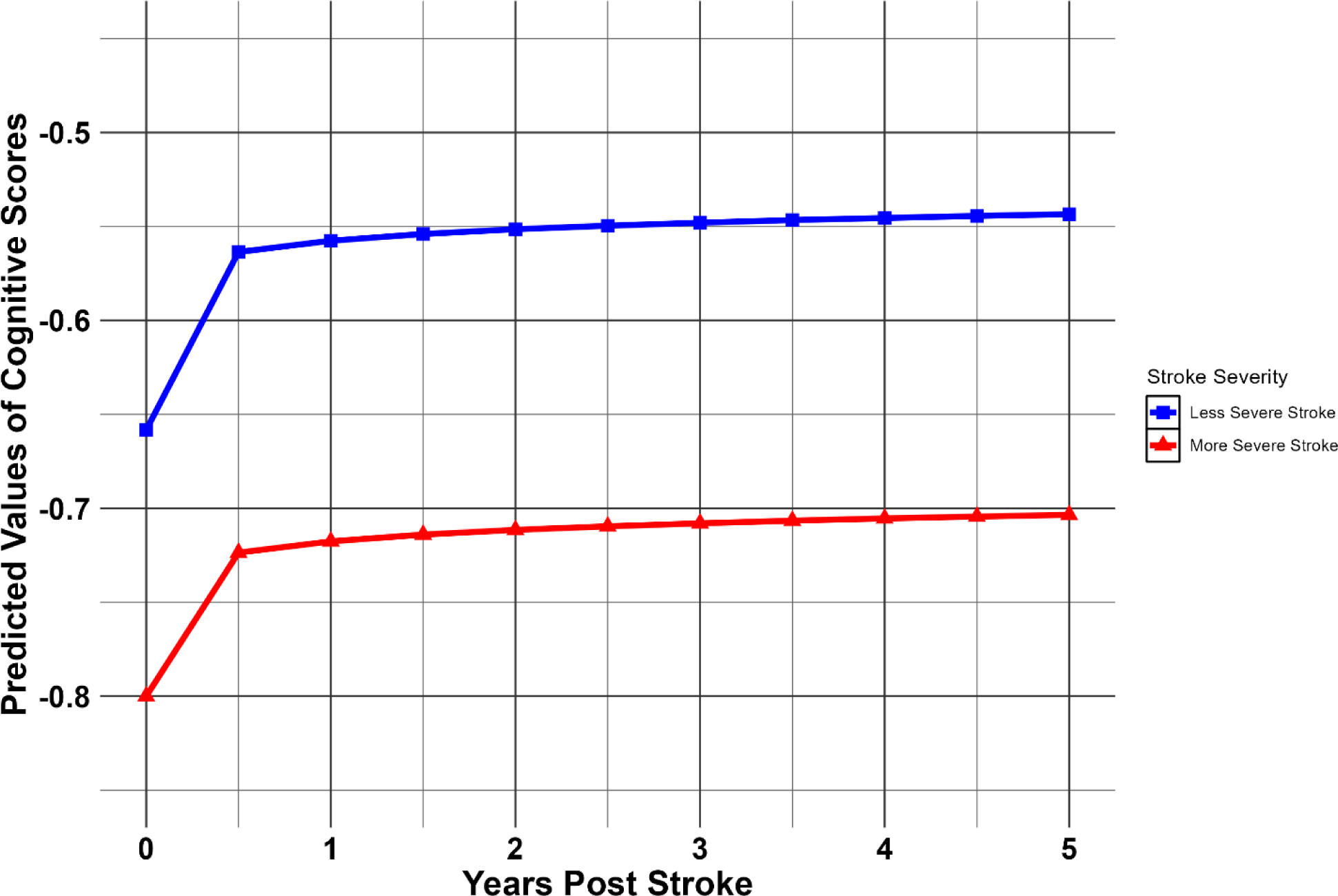
Covariate Adjusted, Logarithmic Trajectories of Cognitive Scores by mRS. The cutoff for ‘less severe stroke’ included any participants with mRS scores less than the median value (1.0). Cutoff for ‘more severe stroke’ was greater than the median value. This model includes baseline cognition, age at enrollment, gender, race, and education as covariates. As this model uses logarithm of time, the first timepoint is estimated at 0.01 years (∼4 days) post stroke.

**Figure 3.**
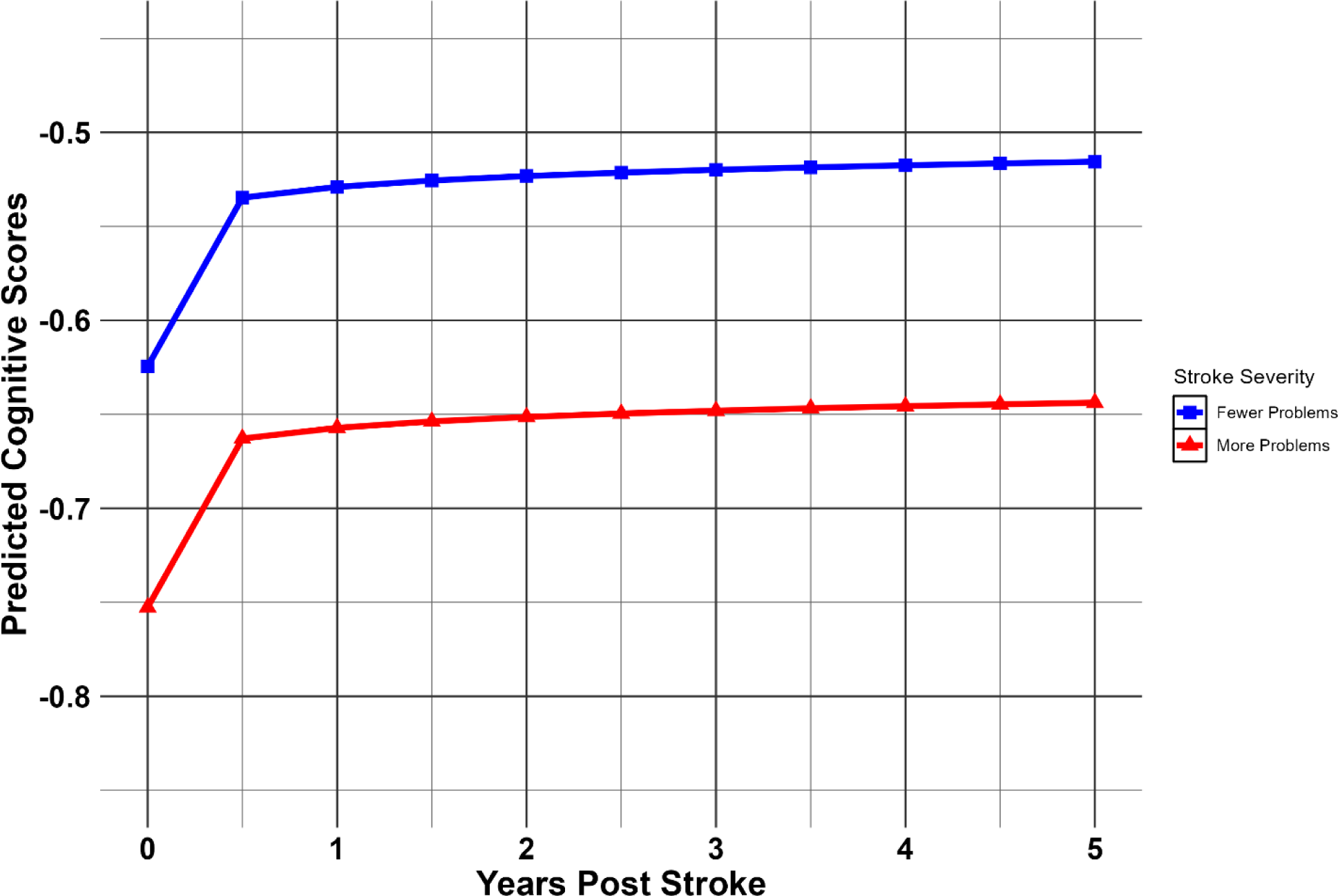
Covariate Adjusted, Logarithmic Trajectories of Cognitive Scores by Caregiver Report. The cutoff for ‘fewer problems’ included all participants whose total number of caregiver-reported problems were less than the median value (6.0). Cutoff for ‘more problems’ include individuals with reported problems greater than or equal to the median value. This model includes baseline cognition, age at enrollment, gender, race, and education as covariates. As this model uses logarithm of time, the first timepoint is estimated at 0.01 years (∼4 days) post stroke.

## RESULTS

The original CARES study recruited approximately 360 stroke survivors initially. However, several exclusions were made to meet the specific criteria for this analysis. Stroke survivors were excluded if the caregiver could not be enrolled (n=31). Dyads were excluded if the stroke survivors did not have both GOS and mRS scores from medical record abstraction (n=78). Additionally, dyads were excluded if the stroke survivors did not have a pre-stroke baseline cognitive assessment (n=42) or if they did not have more than one cognitive assessment over the course of five years following their stroke (n=52). The final sample for this analysis consisted of 157 stroke survivor and caregiver dyads that met these criteria.

**Table 1** displays the demographics of the participants in this sample at the time of the CARES baseline interview, including overall demographics and characteristics of each predictor group. Of the 157 stroke survivors, there was a mean age of 75.74 years (*SD*=7.32) at the time of CARES interview. The mean age at the time of stroke was 74.91 years (*SD*=7.31). The stroke survivor group consisted of 86 women (55%) and included 97 White participants (62%) and 60 Black/African American participants (38%).

**Table 1.** Baseline characteristics of REGARDS-CARES participants.

| *Demographics* | All<br>Participants | High<br>GOS | Low GOS | Low mRS | High mRS | Fewer<br>CGRP | Higher<br>CGRP |
| --- | --- | --- | --- | --- | --- | --- | --- |
|  | N=157 | N=83 | N=74 | N=79 | N=78 | N=73 | N=84 |
| Age at Stroke | 74.91<br>(SD=7.3) | 74.50<br>(SD=7.8) | 75.39<br>(SD=6.7) | 74.22<br>(SD=7.8) | 75.63<br>(SD=6.8) | 74.74<br>(SD=7.5) | 75.07<br>(SD=7.2) |
| Age at CARES<br>interview | 75.77<br>(SD=7.3) | 75.38<br>(SD=7.9) | 76.21<br>(SD=6.7) | 75.11<br>(SD=7.8) | 76.45<br>(SD=6.8) | 75.59<br>(SD=7.5) | 75.93<br>(SD=7.2) |
| Age of Caregiver | 61.82<br>(SD=13.4) | 61.30<br>(SD=14.2) | 62.41<br>(SD=12.5) | 61.53<br>(SD=14.4) | 62.12<br>(SD=12.4) | 66.00<br>(SD=12.2) | 58.23<br>(SD=13.4) |
| Gender (*n*, %) |  |  |  |  |  |  |  |
| Females | 86 (55%) | 42 (51%) | 43 (59%) | 39 (49%) | 47 (60%) | 33 (45%) | 53 (63%) |
| Race (*n*, %) |  |  |  |  |  |  |  |
| White | 97 (62%) | 53 (64%) | 44 (59%) | 51 (65%) | 46 (59%) | 49 (67%) | 48 (57%) |
| Black/AA | 60 (38%) | 30 (36%) | 30 (41%) | 28 (35%) | 32 (41%) | 24 (33%) | 36 (43%) |
| Stroke Type (*n*, %) |  |  |  |  |  |  |  |
| Ischemic | 132 (84%) | 66 (80%) | 66 (89%) | 62 (78%) | 70 (90%) | 63 (86%) | 69 (82%) |
| Hemorrhagic | 13 (8.5%) | 6 (7%) | 7 (10%) | 6 (8%) | 7 (9%) | 5 (7%) | 8 (10%) |
| Not Specified | 12 (7.5%) | 11 (13%) | 1 (1%) | 11 (14%) | 1 (1%) | 5 (7%) | 7 (8%) |
| Stroke Severity |  |  |  |  |  |  |  |
| CGRP<br>at 9 months | 7.09<br>(SD=5.7) | 6.16<br>(SD=5.0) | 8.13<br>(SD=6.3) | 6.18<br>(SD=5.1) | 8.01<br>(SD=6.2) | 2.44<br>(SD=1.7) | 11.13<br>(SD=4.8) |
| GOS<br>at discharge | 4.38<br>(SD=0.7) | 5.0<br>(SD=0.0) | 3.68<br>(SD=0.5) | 5.0<br>(SD=0.0) | 3.74<br>(SD=0.5) | 4.47<br>(SD=0.7) | 4.30<br>(SD=0.8) |
| mRS<br>at discharge | 1.82<br>(SD=1.3) | 0.72<br>(SD=0.6) | 3.05<br>(SD=0.8) | 0.66<br>(SD=0.5) | 3.00<br>(SD=0.8) | 1.56<br>(SD=1.3) | 2.04<br>(SD=1.3) |

*GOS=Glasgow Outcome Scale; mRS=modified Rankin Scale; CGRP=caregiver reported problems*
*Participants were divided into two groups for each predictor based on the 50th percentile (median) score as a cut-off point. The groups of Low GOS, High mRS, and Higher CGRP indicate worse stroke severity. Means and standard deviations are shown for continuous variables. Frequency of categorical variables are shown as a percentage for each group.*

Regarding stroke type, 132 (84%) of the participants experienced an ischemic stroke, 13 (8.5%) had a hemorrhagic stroke, and for 12 (7.5%) participants the stroke type was not specified in the reviewed medical records. Of the 157 caregivers, the mean age at enrollment was 61.82 years (*SD*=13.39), and the majority were female (77%; n=121). The racial composition of the caregiver group paralleled that of the stroke survivor group, with 97 caregivers (62%) identifying as White, 59 (37%) as Black/African American, and one (1%) as “Other.” The mean GOS score was 4.38 (SD=0.72, range = 3 to 5), mean mRS score was 1.82 (*SD=*1.34, range = 0 to 5), and mean number of caregiver-reported problems and impairments was 7.09 (*SD=*5.69, range = 0 to 28).

### GOS and mRS Main Effects

Covariates of age, race, sex, education, baseline scores, and time after stroke were included in all models. In the first model, the GOS score at the time of stroke hospitalization discharge was used as the primary predictor. This model demonstrated a significant main effect of GOS score on cognitive outcomes (b=0.3380, p=0.0009). In the second model, the mRS score at the time of stroke hospitalization discharge was utilized as the predictor variable. This model revealed a significant main effect on cognitive outcomes (b=-0.2119, p=0.0002). There was no main effect of time after stroke on cognitive outcomes in any model included in this analysis.

### Caregiver-Reported Problems Main Effects

The third model included the total number of caregiver-reported problems as the primary predictor variable for cognitive outcomes. In this model, there was a significant main effect of caregiver-reported post-stroke functional ability on cognitive outcomes (b=-0.0671, p<0.0001). The main effect of each separate model and their interaction by time after stroke is displayed in **Table 2**.

**Table 2.** Separate Mixed Models for Each Predictor by Cognition.

| Effects in Longitudinal Model | Estimate (b) | Standard Error | t | p |
| --- | --- | --- | --- | --- |
| GOS | 0.338 | 0.100 | 3.38 | 0.0009 |
| GOS x Time | -0.098 | 0.066 | -1.48 | 0.1408 |
| mRS | -0.212 | 0.055 | -3.89 | 0.0002 |
| mRS x Time | 0.066 | 0.037 | 1.80 | 0.0737 |
| Caregiver-Reported Problems | -0.067 | 0.013 | -5.22 | <.0001 |
| Caregiver-Report x Time | 0.017 | 0.009 | 1.80 | 0.0742 |

*GOS = Glasgow Outcome Scale; mRS = modified Rankin Scale; Time = time of each assessment post-stroke in years*
*This table includes the results from separate mixed models for each predictor. These models are adjusted for the covariates of baseline cognition, time after stroke, age, sex, race, and education. Low GOS scores, high mRS scores, and higher numbers of Caregiver-Reported Problems indicate worse stroke severity*

### Comparing Predictors

A final model included all three predictors and covariates to assess the contribution of each predictor, as well as all previously included covariates. When all predictors were considered simultaneously, caregiver-reported problems continued to be a significant predictor of cognitive outcomes (b=-0.0480, p<0.0001). Neither GOS (b=0.02511, p=0.8742) nor mRS (b=-0.1079, p=0.2216) showed a statistically significant effect on cognitive outcomes in the presence of other predictors.

Standardized beta coefficients for the three predictors were also calculated to provide insight into their relationship to longitudinal cognitive scores. The standardized betas for the predictors are as follows: caregiver report (*B*=-0.4341), GOS (*B*=0.2773), and mRS (*B*=-0.3250). These were calculated using the mean and standard deviation of each predictor and the pre-stroke baseline cognitive score (*M*=-0.2669, *SD*=0.8790).

## DISCUSSION

The current investigation revealed significant main effects of all three predictors on longitudinal cognitive outcomes. The positive relationship between GOS and cognition indicates that a higher GOS score, representing better acute functioning, is associated with a higher cognitive score over time. Conversely, the negative relationship between mRS and cognitive composites demonstrates that as mRS scores increase, representing worse stroke-related disability, cognitive scores tend to decrease. This confirms the study hypothesis that more severe stroke, as measured by acute outcome scales, is predictive of lower cognitive scores in stroke survivors. This finding supports the current literature and demonstrates that clinically administered measures of stroke severity are prognostic of longitudinal cognitive scores in stroke survivors, emphasizing the importance of considering acute phase functioning measures when evaluating cognitive recovery.^48, 49^

This analysis also supported the link between caregiver-reported functioning and stroke survivor cognition over time. Specifically, higher numbers of caregiver-reported problems were associated with lower cognitive composite scores over time. These findings support the study hypotheses and indicate that both caregiver reports and acute clinical measures have value in predicting cognitive outcomes in stroke survivors. The findings also align with previous research on the critical role of caregivers in understanding and monitoring care recipient outcomes.^50, 51^ In all models, there was a significant main effect of baseline composite scores on cognitive outcomes. This finding is expected, as pre-stroke cognitive function is known to precipitate cognitive outcomes after stroke.^52^

A comparison of the absolute values of the standardized betas revealed the relative strength of each predictor. Amongst the three predictors, the caregiver-report of functional impairment had the strongest association with cognitive outcomes, followed by the mRS scores and then GOS scores. These results suggest that assessing functional impairment is crucial for understanding cognitive outcomes in stroke survivors. Notably, caregiver-reported problems demonstrated the strongest relationship with cognitive outcomes, emphasizing the importance of incorporating caregivers’ perspectives in stroke assessment and recovery monitoring.

The strong association between number of caregiver-reported problems and cognitive scores may be explained by several factors. Caregiver-reported measures provide a more comprehensive assessment of the impact of stroke on daily functioning, including problems with ADLs and IADLs, which are significantly affected by impairments cognitive abilities. Also, it is important to note that the caregiver-reported problems were collected an average of nine months after the stroke event, while the other two measures were collected from the medical records at the time of stroke hospitalization discharge. This information is especially valuable as it comes after the six-month critical recovery window, a time when substantial functional changes can still occur.^53^ Therefore, incorporating caregiver-reported measures at later stages can provide a more comprehensive and nuanced understanding of cognitive outcomes in stroke survivors.

The implications of these findings provide further evidence that integrating caregiver-reported problems with clinical measures can provide a more comprehensive understanding of stroke survivors’ cognitive recovery. While a brief informant report would not replace post-stroke clinical follow-up visits, they often support clinical decision making to evaluate changes in cognition and better address the specific needs of each patient. This is especially prevalent in patients with cognitive impairment where patient’s often suffer from lack of awareness of deficits (cognitive anosognosia).^54^ Cognitive anosognosia is common in mild cognitive impairment patients and makes informant assessments crucial to a post-stroke dementia diagnosis.^24, 27^

These results also emphasize the importance of monitoring stroke survivors longitudinally using telephone cognitive assessments. Regular assessments following hospitalization can monitor the cognition of stroke survivors while also capturing the dynamic relationship between caregiver-reported problems and cognitive outcomes. Collecting caregiver reports is also cost-effective and time-efficient compared to other assessments, as caregivers can provide expedited updates without the need for repeated clinical visits, especially in a population that commonly has limited insight into their own functioning. Incorporating caregiver reports of stroke impairment into the assessment of stroke survivors provides valuable information for clinicians and researchers designing interventions.

This analysis has several strengths, such as the inclusion of multiple longitudinal assessments for many of the stroke survivors. These longitudinal assessments provide a comprehensive insight into the time-dependent relationship of stroke severity and recovery. Another strength is the inclusion of pre-stroke cognitive scores as well as demographic variables as covariates that ensure a rigorous control for potential confounding variables. Also, the use of a community-based sample enhances the generalizability of the findings due to the diverse and large sample size.

A limitation of this analysis is that it relies on secondary data, as the original research design was not specifically tailored to address the research questions of this analysis. Therefore, this analysis was limited to utilizing the mRS and GOS only and did not include other common measures of stroke outcomes and severity, such as the NIHSS. Also, the original CARES study involved purely observational methods. Therefore, the conclusions that can be drawn are correlative in nature only and causation of effects cannot be determined through analysis of this study data. A potential source of bias stems from the subjectivity of the caregiver-reported measure of post-stroke functioning. However, this is counterbalanced by the inclusion of the outcome measures from hospitalization discharge along with the caregiver-reported problems. These collectively provide a more comprehensive evaluation of stroke impairment than clinical assessments alone.^24^ The timing of the caregiver-report allowed for a more detailed assessment of stroke-related impairments compared to the post-hospitalization measures that are collected in the acute phase and therefore outside of the critical recovery window.

Future reports from this analysis will seek to understand factors that moderate the relationship between stroke severity and cognition. Future investigations should also consider the multidirectional relationship between the caregiver’s experience and the patient’s cognitive outcomes. Additionally, these studies should examine the longitudinal relationship between caregiver-reported impairments, acute phase stroke severity measures, and longitudinal cognitive outcomes in a larger, more diverse sample of stroke survivors.

## Conclusion

This study highlights the critical role of caregiver-reported impairments, alongside other stroke severity measures, in predicting cognitive outcomes in stroke survivors. These findings highlight the value of integrating both clinical assessments and informant perspectives in order to contribute more accurate detection of post-stroke cognitive impairment and dementia. Utilizing caregiver reports of everyday function can also improve prediction of cognitive outcomes and facilitate tailoring interventions to the evolving needs of stroke survivors. Ultimately, this analysis enhances the understanding of the complex interplay of factors influencing cognitive impairment, reaffirming the need for ongoing assessments and caregiver involvement in understanding the cognitive consequences of stroke.

## Data Availability

REGARDS has policies and procedures currently in place to permit access to study data for manuscripts through a review and approval process under the governance of the REGARDS Executive Committee. The corresponding author does not determine how to make the data in this manuscript available. Please contact to inquire and request access to the data referred to in the manuscript.

## Acknowledgements

This research project is supported by cooperative agreement U01 NS041588 co-funded by the National Institute of Neurological Disorders and Stroke (NINDS) and the National Institute on Aging (NIA), National Institutes of Health, Department of Health and Human Service. The content is solely the responsibility of the authors and does not necessarily represent the official views of the NINDS or the NIA. Representatives of the NINDS were involved in the review of the manuscript but were not directly involved in the collection, management, analysis, or interpretation of the data. The authors thank the other investigators, the staff, and the participants of the REGARDS study for their valuable contributions. A full list of participating REGARDS investigators and institutions can be found at: https://www.uab.edu/soph/regardsstudy/

## Author Contributions

D.L.R. & O.J.C. designed and implemented the original experiment. J.A.B. conceptualized this manuscript, performed the data analysis, and interpreted the data. Analysis was assisted primarily by D.L.L., S.E.J. & O.J.C.

J.A.B. wrote the manuscript, all additional authors provided supporting conceptualization and manuscript revisions. D.L.R. secured partial funding for the initial CARES ancillary study.

